## Supplemental materials for "Protection of COVID-19 vaccination and previous infection against Omicron BA.1, BA.2 and Delta SARS-CoV-2 infections"

**Supplementary material**

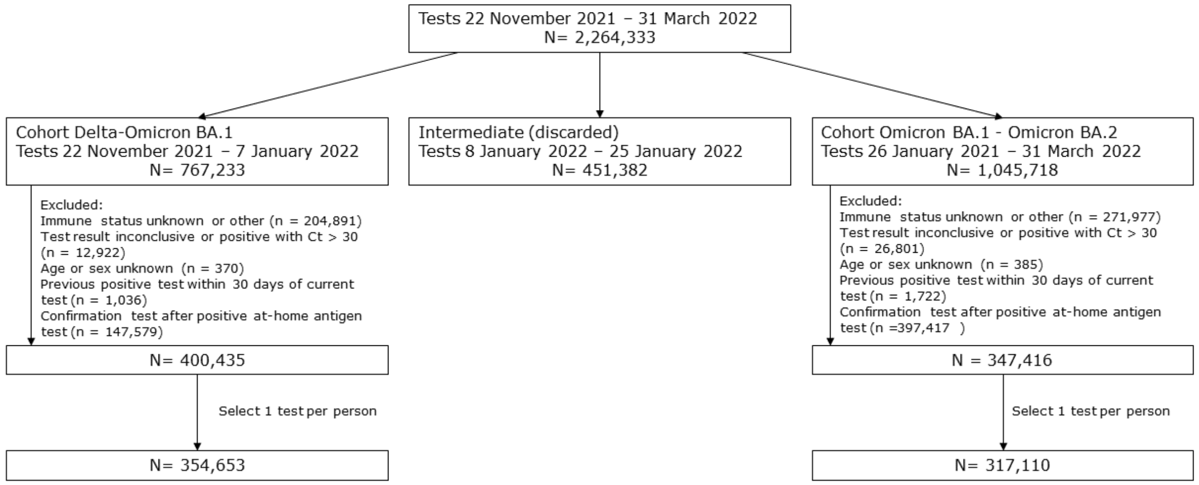

Figure S1. Flowchart of data selection

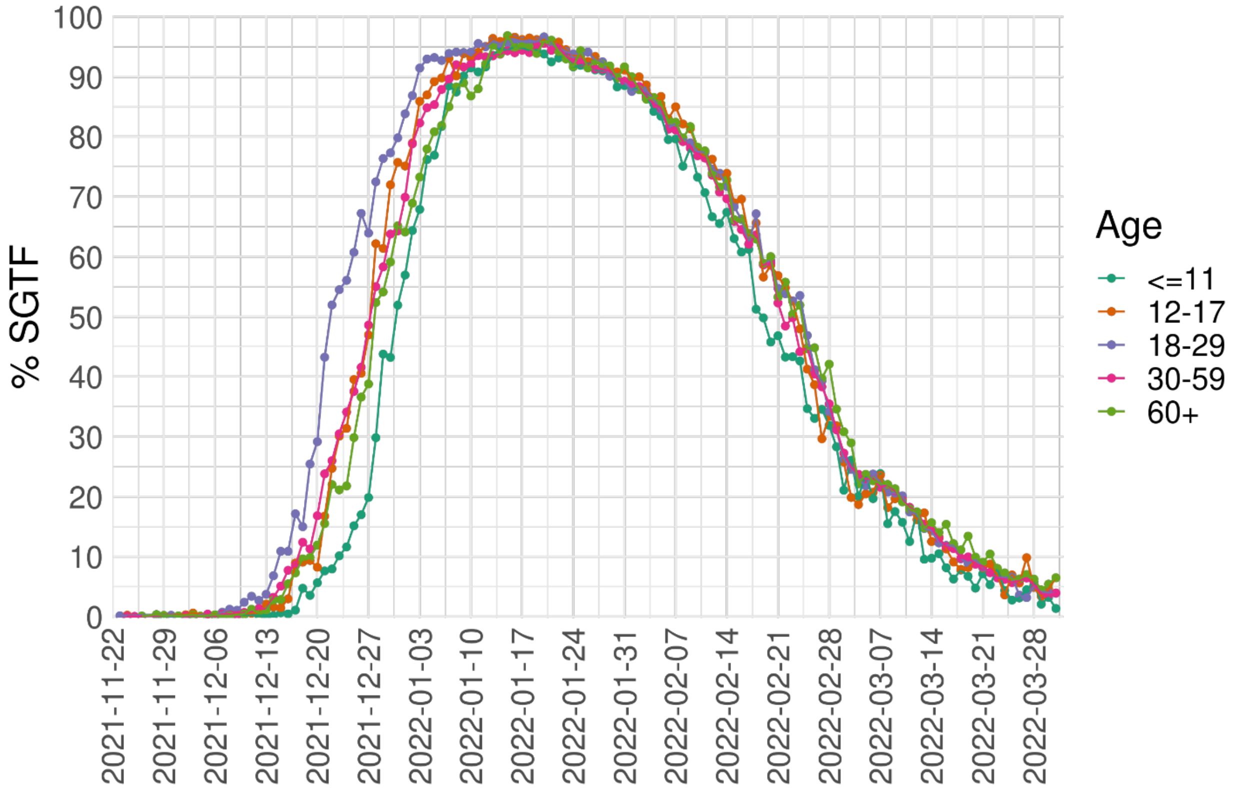

Figure S2. Proportion of SGTF among positive SARS-CoV-2 cases by age group, the Netherlands, 22 November 2021 - 31 March 2022, n = 941,692.

Table S1 WGS variant found in SGTF (S result, not detected) and non-SGTF (S result, detected) cases in each cohort. First column indicates the cohort, followed by the SGTF result. The number of WGS typed Omicron BA.1, Delta and BA.2 cases. Row-wise calculations on positive predictive value (PPV) and column-wise calculations on sensitivity.

| Cohort | S result | WGS result Omicron BA.1 | WGS result Delta | WGS result Omicron BA.2 | PPV | Sensitivity |
|---|---|---|---|---|---|---|
| Cohort Delta-Omicron BA.1 | Not detected | 132 | 0 | 0 | 1.00 | 1.00 |
| Cohort Delta-Omicron BA.1 | Detected | 0 | 353 | 0 | 1.00 | 1.00 |
| Cohort Omicron BA.1-BA.2 | Not detected | 158 | 0 | 0 | 1.00 | 0.99 |
| Cohort Omicron BA.1-BA.2 | Detected | 2 | 0 | 128 | 0.98 | 1.00 |

Table S2 Relative reduction in Delta and Omicron BA.1 infections after previous infection, primary vaccination, booster vaccination, or combinations of previous infection and vaccination (column 'Vaccination_and_previous_infection_status'), compared with naïve status ((1-OR) * 100), by time since last event and overall (column 'Time_since_event') in persons aged 18 and by variant (column 'variant'), for cohort Delta-Omicron BA.1 and cohort Omicron BA.1-BA.2 (column 'Cohort'). Estimates and 95%-CI (in brackets) are calculated for all cases (column 'VE_CI') and cases with reported symptoms (column 'VE_CI_symptomatic_infection').

Table S3 Protection estimates for booster vaccination and booster vaccination with a previous infection with 'Primary vaccinated' as reference.

| Cohort | Immuunstatus | Variant | VE_CI |
|---|---|---|---|
| Delta-Omicron BA.1 | Booster | Omicron BA.1 | 42% (38-46) |
| Delta-Omicron BA.1 | Booster | Delta | 73% (70-77) |
| Delta-Omicron BA.1 | Previous infection, booster | Omicron BA.1 | 53% (45-61) |
| Delta-Omicron BA.1 | Previous infection, booster | Delta | 92% (83-96) |
| Omicron BA.1-BA.2 | Booster | Omicron BA.1 | 47% (45-48) |
| Omicron BA.1-BA.2 | Booster | Omicron BA.2 | 40% (38-42) |
| Omicron BA.1-BA.2 | Previous infection, booster | Omicron BA.1 | 68% (66-70) |
| Omicron BA.1-BA.2 | Previous infection, booster | Omicron BA.2 | 67% (65-69) |

Table S4 Relative reduction in Delta and Omicron BA.1 infections after previous infection, primary vaccination, booster vaccination, or combinations of previous infection and vaccination (column Vaccination_and_previous_infection_status), compared with naïve status ((1-OR) * 100), by time since last event and overall (column Time_since_event), by age group (column Age_group) and by variant (column 'variant'), for cohort Delta-Omicron BA.1 and cohort Omicron BA.1-BA.2 (column Cohort).

Table S5 Interaction terms between age and vaccination and previous infection status per cohort and variant with age group 18-29 years old as reference group.
